# Balancing optimization and standardization in multisite fMRI data analyses to address site-specific parameters

**DOI:** 10.64898/2026.09.04.26362236

**Authors:** Marie-Eve Hoeppli, Saül Pascual-Diaz, Emma E. Biggs, Laura Simons, Robert C. Coghill, Marina López-Solà, Christopher D. King, Nima Aghaeepour, Martin Angst, Brice Gaudilliere, Jennifer Stinson, Massieh Moayedi

## Abstract

Multisite functional Magnetic Resonance Imaging (fMRI) studies are rapidly becoming the norm to allow the acquisition of large samples required to perform advanced statistical techniques, e.g. machine learning. However, acquiring data at multiple sites includes methodological and technical challenges due to the difference in the facilities of each site, such as scanner manufacturer and model. These new challenges add to the already well-known challenges of fMRI data, including scanner-related noise, participant movement, and physiological confounds. To ensure the optimal quality of data, a balance needs to be achieved between standardizing data preprocessing across sites and optimizing within-site data preprocessing. To define the optimal preprocessing pipeline including standard steps and additional denoising technique for our multisite dataset, we first tested 3 commonly used preprocessing pipelines, i.e. fMRIPrep, FSL, and CONN. These pipelines all include standard steps, e.g. motion correction, temporal filter, etc. In a second step, to further improve signal quality, we tested the efficiency of 3 additional denoising techniques on the output of the data preprocessed with the previously defined pipeline. These techniques included aCompCor, FSL FIX and ICA-AROMA. Signal quality was quantified as temporal signal-to-noise ratio (tSNR) across and within sites. Our results show that the performance of the preprocessing pipeline and denoising technique varies between sites. FSL yielded the highest tSNR at two sites and fMRIPrep at the third, revealing a significant site x pipeline interaction. An FSL-based pipeline with minimal adjustment to accommodate site-specific parameters and followed by denoising using a single FSL FIX classifier, which was custom-trained across sites, achieved the highest quality of signal in our data and yielded the greatest consistency in improved data quality across sites. Because of the potential of this pipeline to adequately identify and address site-specific noise, while remaining constant across sites, we selected it as optimal preprocessing pipeline for our dataset.

## 1. Introduction

In recent years, there has been a significant increase in the collection of multisite data in functional Magnetic Resonance Imaging (fMRI) research. The ability to collect large samples, particularly when studying patient populations that are challenging to recruit ^1^, has become increasingly important for taking advantage of the advanced statistical techniques, e.g. machine-learning algorithms ^2^, that are rapidly becoming the new norm for fMRI analyses. These techniques are increasingly valuable in advancing our understanding of complex neural mechanisms, such as the ones involved in chronic pain. Multisite designs also facilitate collaboration across imaging centers and increase sample diversity, thereby improving the generalizability of findings beyond the population and scanner characteristics of a single center.

Despite these advantages, the integration of multisite data poses substantial methodological and experimental challenges ^1^. Some challenges are similar to those encountered in single-site studies, e.g. the need to remove scanner-related, motion-related, and physiological noise while retaining the signal of interest. Due to the noisy nature of fMRI data, optimal preprocessing is essential to retain the signal of interest while removing the noise ^3,4^. Although the importance of good signal quality is well-known throughout the neuroimaging community, the importance of optimized fMRI data preprocessing is often underestimated and underinvestigated. Suboptimal preprocessing and denoising pipelines might complicate signal extraction and even raise questions about the validity of the findings.

Other challenges are specific to multisite designs. Even when the same pulse sequences are programmed at every site, differences in scanner manufacturer, hardware, software, and procedures required at each imaging center, introduce site-specific noise characteristics. These differences may also constrain acquisition parameters, e.g. vendor-specific implementation of multiband acceleration or online distortion correction. These site-specific effects introduce complexities in the preprocessing and analysis of the collected data. Consequently, a given preprocessing step may not perform equally well on data from different scanners, and some steps may be required at one site but not another. Such differences directly affect the comparability of the multisite data.

Our current study, in which we aim to investigate predictors of recovery in pediatric chronic musculoskeletal pain, includes two additional challenges. First, pain is associated with a high level of physiological noise, whose removal requires to strike a fine balance between noise reduction and signal loss ^5^. Second, children have previously been shown to exhibit greater physiological noise than adults, due to normal physiological differences between the two, e.g. higher heart rate in children, and developmental processes ^6^. Pediatric pain populations therefore represent a demanding test case for any preprocessing pipeline.

Addressing all these challenges raises a question that is rarely made explicit in multisite studies: should the goal of preprocessing be standardization (i.e. applying an identical pipeline to all sites so that data is comparable), or site-specific optimization (i.e. adapting the pipeline to each site to maximize data quality)? Standardization is crucial to ensure comparability and to minimize the impact of confounding factors that can be detected across sites, allowing for more robust and reliable analyses. Nevertheless, standardization by itself does not remove site-specific noise, which is then carried forward into the analyses. On the other hand, optimizing the processing procedures for each site dataset avoids biasing the results towards any site, reducing site-specific noise, and capitalizes on the unique strengths of each dataset. Previous discussions surrounding multisite dataset processing have primarily focused on harmonization by employing second-level statistical techniques to control for potential site-specific effects and establishing standard approaches to data collection and analysis across sites ^7–12^. While this harmonization approach has yielded valuable insights to ensure the comparability of the data, this one-size-fits-all approach is likely insufficient for achieving optimal fMRI data quality. Taking into consideration site-specific needs is essential for this purpose. Depending on scanner settings, different sites might require different preprocessing steps. Within-site optimization of the preprocessing procedures also reduces the noise specific to a site, further optimizing the quality of the signal available for analyses.

Here, we first aim to highlight the main challenges encountered when working with multisite fMRI. Second, we aim to define possible approaches to effectively balance the competing priorities of standardization across sites and optimization of individual site datasets. By acknowledging the challenges and exploring potential solutions, we seek to contribute to the ongoing efforts in improving the quality and reliability of multisite fMRI data analyses.

## 2. Methods

### 2.1. Study design

The data presented in this paper was collected as part of a multisite prognostic biomarker study ^13^. Three sites were included in this study (Stanford University, Cincinnati Children’s Hospital, and SickKids Hospital Toronto). The imaging center at each site was equipped with a different scanner (GE at Stanford, Philips in Cincinnati, and Siemens in Toronto), which might be responsible for differences in the dataset. In addition, the differences in scanner might require site-specific adjustments in data processing to address the specificity of each scanner. Details about the experimental design and data management are reported elsewhere ^13^. To define the optimal preprocessing pipeline for the preparation of the main study’s resting-state fMRI data for analyses, a subset of data acquired in this study was selected. Two steps were defined to optimize the pipeline to prepare our data for analyses: (1) defining the optimal preprocessing pipeline for our data following standard preprocessing steps; (2) evaluating the value of adding a denoising technique to the previously defined pipeline and defining its efficiency. At each step, the temporal signal-to-noise ratio (tSNR) was compared between preprocessing pipelines, respectively denoising techniques, within and across regions of interest (see section 2.4).

In the first step, we compared three well-known preprocessing pipelines including standard steps to prepare data for analyses: fMRIPrep ^14,15^, an FSL-based preprocessing pipeline ^16–18^, and CONN ^19^ (Figure 1). In this first step, we also evaluated the effect of using high-pass temporal filtering or bandpass temporal filtering during the preprocessing. In the second step, after defining the best performing preprocessing pipeline, i.e. FSL with bandpass temporal filtering, we evaluated the added benefit of three denoising algorithms: aCompCor ^20^, FIX ^21,22^, and ICA-AROMA ^23^ (Figure 2).

**Figure 1.**
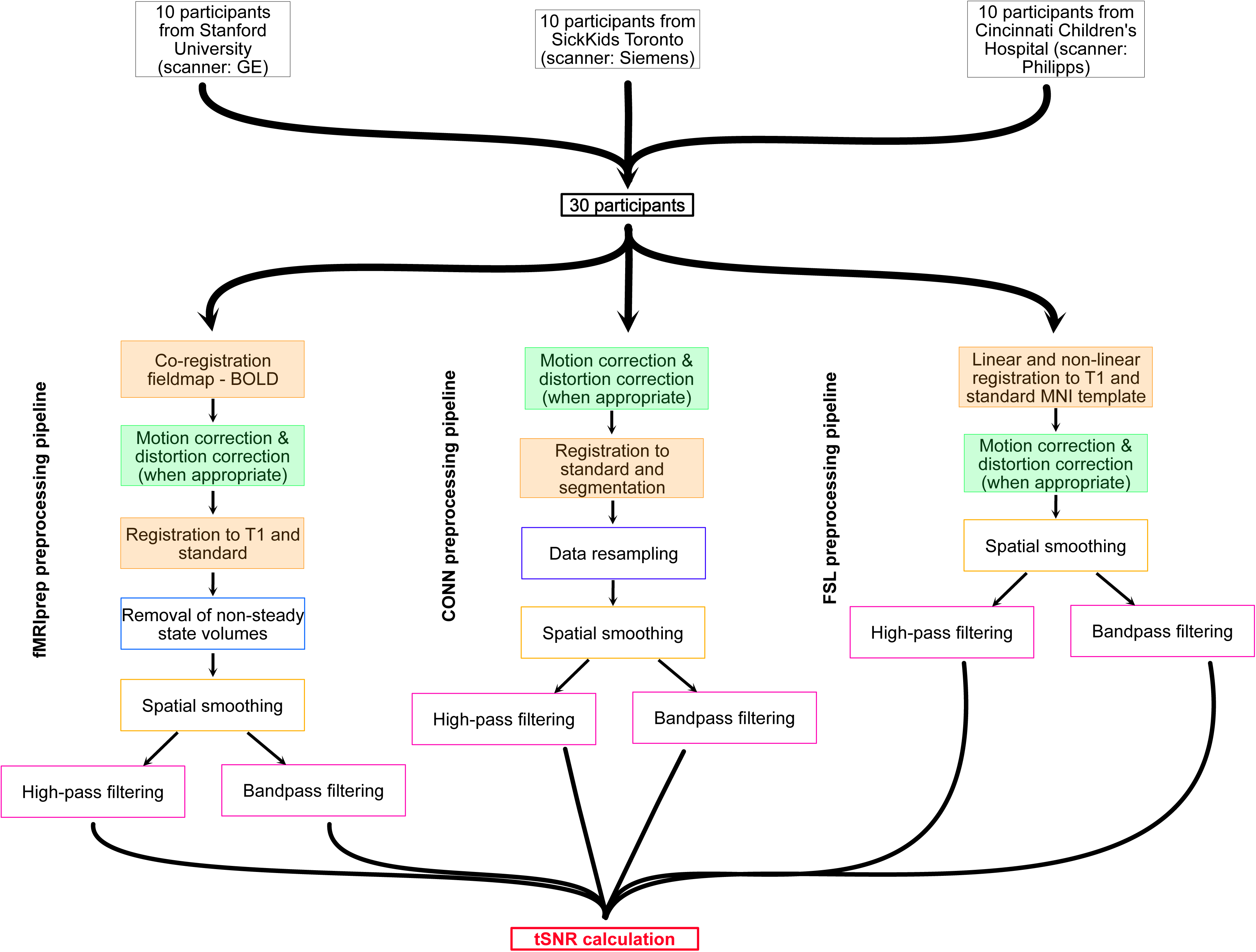
Flow of the three preprocessing pipelines tested in this study including their respective preprocessing steps. Ten participants were selected at each of the three sites to investigate the effect of the acquisition site on the efficiency of the preprocessing. Two steps, i.e. registration and motion correction/unwarping, were manually inspected (filled rectangle). Preprocessed data was then used for tSNR calculation.

**Figure 2.**
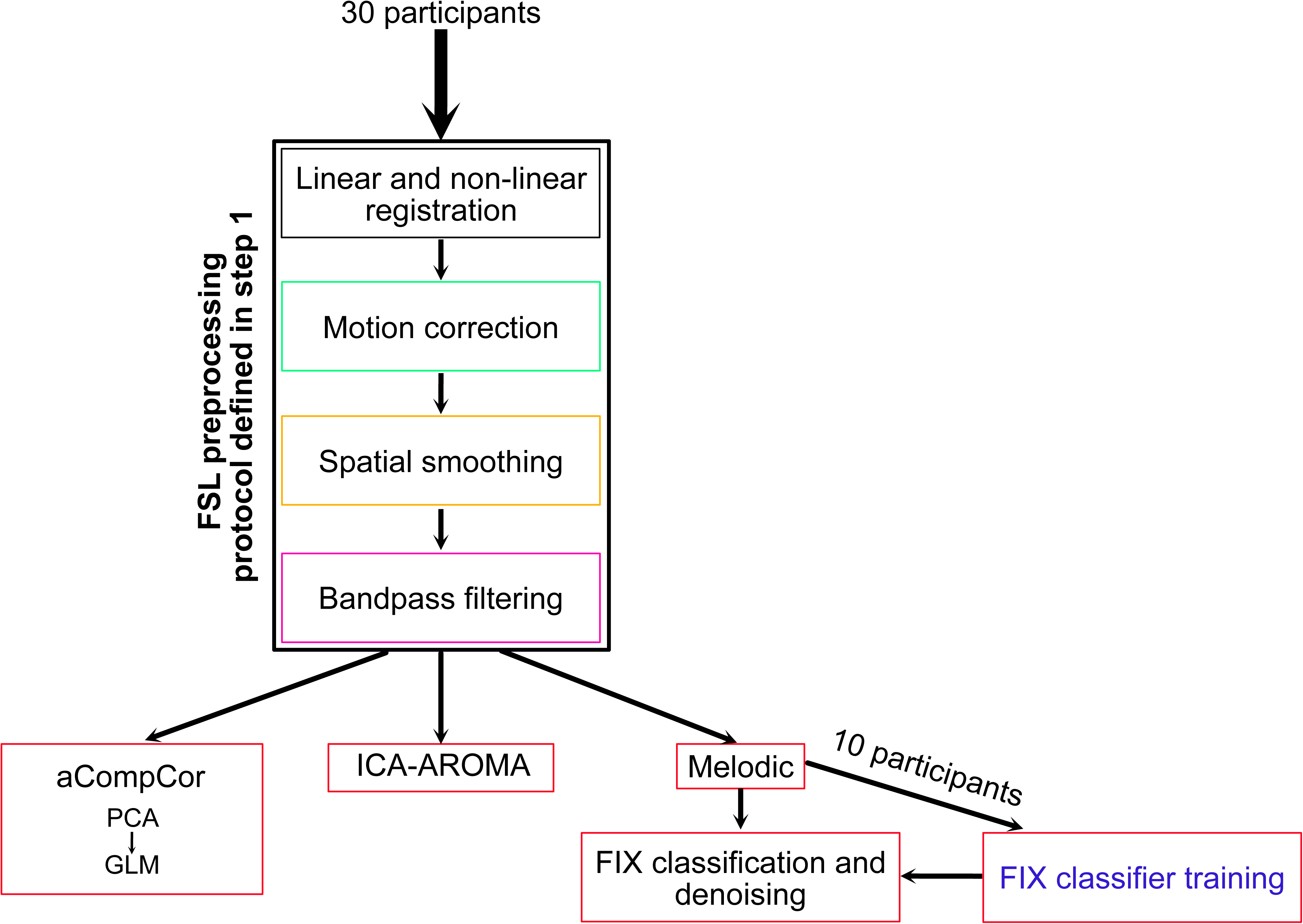
Flow of the denoising techniques tested in this study. Denoising techniques were applied to the data preprocessed with the pipeline previously defined as optimal, i.e. FSL with bandpass filter. PCA: Principal Component Analysis; GLM: General Linear Model.

At both steps, to evaluate the efficacy of the pipeline in improving the quality of the signal, manual inspection of the preprocessing steps, including the inspection of the motion correction and registration/standardization, was used, and tSNR was calculated and statistically compared between pipelines. tSNR is a well-recognized measure of the quality of the signal in a dataset. Manual inspection was performed after motion correction/unwarping and after registration (figure 1), before the tSNR extraction

### 2.2. Participants

Ten adolescents diagnosed with chronic musculoskeletal pain condition were selected at each site (Table 1), resulting in 30 participants across sites. Among these, at each site, 5 of the 10 chosen participants exhibited a high level of noise in their raw resting-state data and 5 participants exhibited minimal noise. The level of noise was defined through manual inspection of the motion and artifacts observed in the raw data. This was to ensure that the chosen preprocessing pipeline was efficient regardless of how noisy the data was. All participants met the eligibility criteria and completed the whole study as described in Simons et al. ^13^.

**Table 1.** Demographics.

| SITE | AGE<br>(MEAN ± SD) | SEX (MALE; FEMALE) |
| --- | --- | --- |
| CINCINNATI | 15.0 ± 2.1 | 1; 9 |
| STANFORD | 16.1 ± 1.1 | 1; 9 |
| TORONTO | 15.0 ± 1.8 | 2; 8 |
| OVERALL | 15.4 ± 1.7 | 4; 26 |

### 2.3. Data acquisition parameters

Data was acquired on 3T scanners from three manufacturers (GE, Philips, Siemens) at three different sites (The Richard M. Lucas Center for Imaging at Stanford School of Medicine, Cincinnati Children’s Hospital Medical Center, and SickKids Hospital Toronto; Table 2).

**Table 2.**
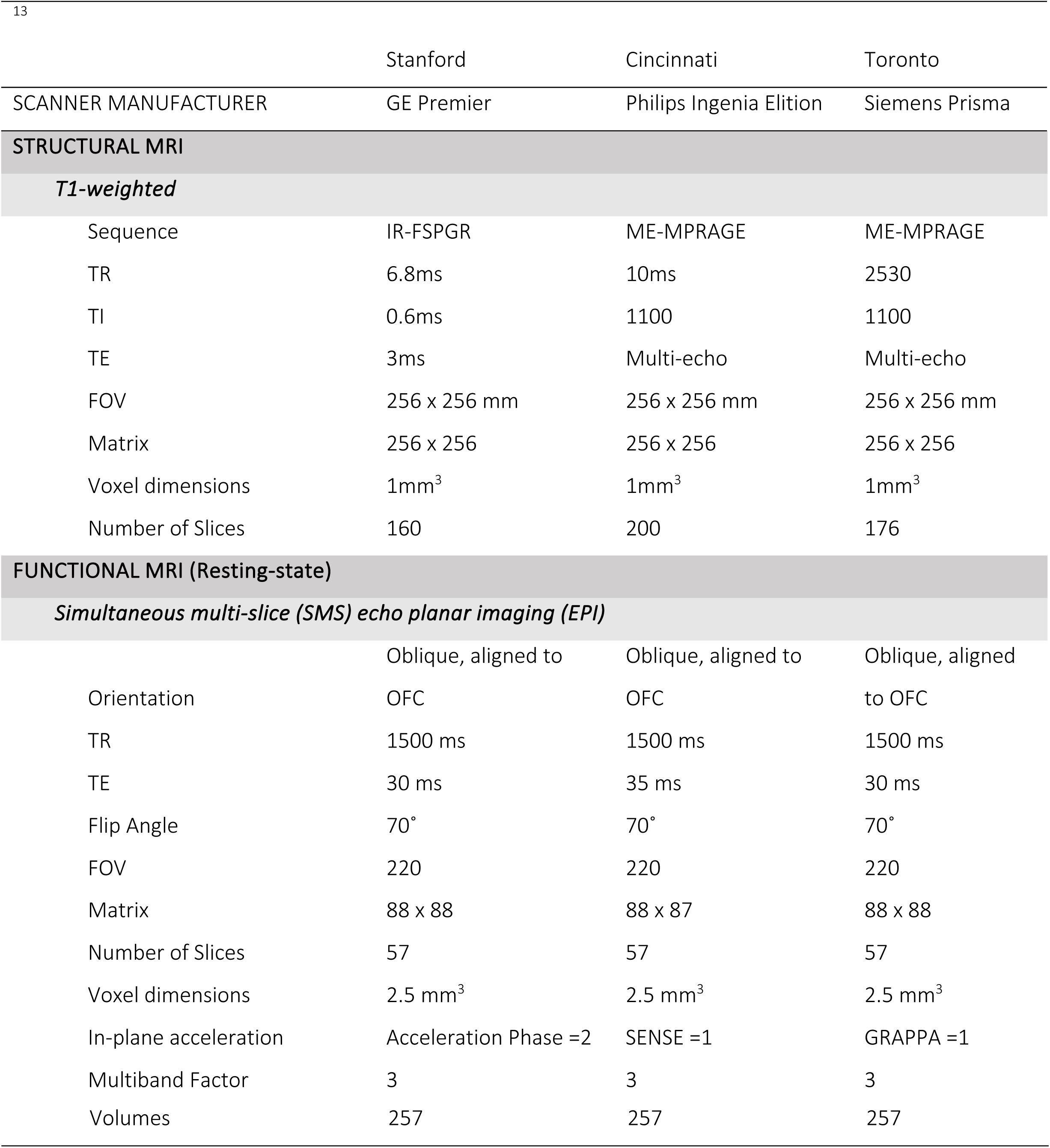
Acquisition parameters at each site. ^13^

### 2.4. Data quality metrics and regions-of-interest

To evaluate the quality of each preprocessing pipeline on the data, qualitative and quantitative inspection of the preprocessed data was performed. The qualitative inspection included the visual inspection of the motion correction parameters and unwarping (if applicable), as well as the visual inspection of the registration and standardization. The evaluation criteria were defined in advance and in accordance with MRIQC guidelines (https://mriqc.readthedocs.io/en/latest/index.html,^24^) (Table 3).

**Table 3.** Evaluation criteria used for the visual inspection of the motion correction, unwarping, and registration.

|  | **Registration** | **Unwarping** | **Motion** |
| --- | --- | --- | --- |
| **good** | All boundaries aligned | Improved fit/no change | <2mm |
| **maybe** | GM/WM boundary correct | Over-correction cerebellum | 2-4mm |
| **bad** | Incorrect alignment | Shearing, distortions | >4mm |

For the quantitative evaluation of the effect of the preprocessing pipeline on the data, head motion parameters were extracted from the motion correction preprocessing step and tSNR was extracted from regions of interest (ROIs) that are relevant to the studied population. All tSNR maps were computed in the MNI space (2 mm isotropic) after the final spatial normalization step of each pipeline, so that identical ROI masks could be applied across pipelines. Head motion was summarized as mean framewise displacement (FD) computed from the six rigid-body motion parameters (three translations in mm, three rotations converted to mm).

tSNR is a common measure of signal quality ^25,26^. It was calculated by dividing the mean signal over time by the standard deviation of the signal over time (equation 1).

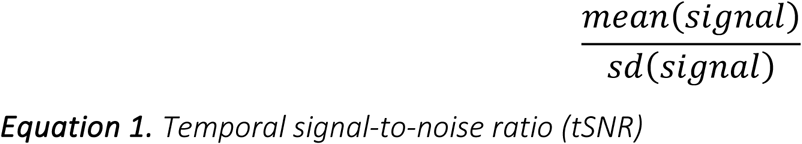

### Equation 1. Temporal signal-to-noise ratio (tSNR)

Given the focus of the main study on predictors of chronic pain, ROIs were selected based on previous literature and included brain areas previously associated with chronic pain or regions known to be associated with high level of physiological noise and little to no signal of interest, e.g. cerebrospinal fluid (CSF) and white matter (WM). The amygdala, insula, ventromedial prefrontal cortex (vmPFC), orbitofrontal cortex (OFC), anterior cingulate cortex (ACC), midcingulate cortex (MCC), posterior cingulate/precuneus (PCC), hippocampus, and thalamus ROIs were defined from the Harvard-Oxford (sub-)cortical atlas ^27^. The primary (S1) and secondary (S2) somatosensory cortices were defined from the Juelich Histological atlas ^28^ and WM and CSF masks were created from SPM tissue probability maps (TPM) (https://www.fil.ion.ucl.ac.uk/spm/). ACC-MCC corresponds to the Harvard-Oxford “cingulate gyrus, anterior division” parcel, yielding 10 ROIs for the statistical analyses.

### 2.5. Preprocessing

We applied the following preprocessing steps (when appropriate) using three popular toolboxes (fMRIPrep, FSL, and CONN/SPM): intensity inhomogeneity correction and skull stripping of T1 images, motion correction, temporal filtering using high-pass (0.008 Hz) or bandpass filtering (0.008 Hz – 0.09 Hz), spatial smoothing (6mm FWHM Gaussian kernel in all three pipelines), co-registration, unwarping (for data from Stanford and Toronto sites), and standardization of T2* images. Data from the Cincinnati site underwent online unwarping during data reconstruction; thus, no further unwarping was applied on these data to prevent overcorrection. Toolbox-specific steps are described below; parameters that could be matched across toolboxes (temporal filters, smoothing kernel, and output space) were held constant.

#### 2.5.1. Preprocessing in fMRIPrep

Results included in this manuscript come from preprocessing performed using fMRIPrep 24.1.1 ^14,29^, which is based on Nipype 1.8.6 ^30^. The T1-weighted (T1w) images were corrected for intensity non-uniformity (INU) with N4 Bias Correction ^31^, distributed with ANTs 2.3.3 ^32^, and used as T1w-reference throughout the workflow. The T1w-reference was then skull-stripped with a Nipype implementation of the antsBrainExtraction.sh workflow (from ANTs), using OASIS30ANTs as target template. Brain surfaces were reconstructed using recon-all (FreeSurfer 7.2), and brain tissue segmentation of CSF, WM, and gray-matter (GM) was performed on the brain-extracted T1w using FAST (FSL 5.0.9, ^33^). Volume-based spatial normalization to two standard spaces was performed through nonlinear registration with antsRegistration (ANTs 2.3.3), using brain-extracted versions of both T1w reference and the T1w template.

For the BOLD runs, the following preprocessing was performed. First, a reference volume and its skull-stripped version were generated using a custom methodology of fMRIPrep. Skull stripping was achieved using BET, FSL brain extraction tool^34^. At Toronto and Stanford sites, a B0-nonuniformity map (or fieldmap) was directly measured with an MRI scheme designed with that purpose. The fieldmap was then co-registered to the target echo-planar imaging (EPI) reference run and converted to a displacement fieldmap (amenable to registration tools such as ANTs) with FSL’s fugue and other SDCflows tools. Based on the estimated susceptibility distortion, a corrected EPI reference was calculated for a more accurate co-registration with the anatomical reference. The BOLD reference was then co-registered to the T1w reference using FLIRT (FSL 5.0.9) ^35^ with the boundary-based registration ^36^ cost function. Co-registration was configured with nine degrees of freedom to account for distortions remaining in the BOLD reference. Head-motion parameters with respect to the BOLD reference (transformation matrices, and six corresponding rotation and translation parameters) were estimated before any spatiotemporal filtering using MCFLIRT (FSL 5.0.9) ^37^. The BOLD time-series were resampled onto their original, native space by applying a single, composite transform to correct for head-motion and susceptibility distortions. These resampled BOLD time-series will be referred to as preprocessed BOLD in original space, or just preprocessed BOLD. The BOLD time-series were resampled into standard space, generating a preprocessed BOLD run in MNI152NLin6Asym space (2 mm isotropic). Non-steady state volumes were removed and spatial smoothing with an isotropic, Gaussian kernel of 6mm FWHM (full-width half-maximum) was applied to the data. Several confounding time-series were calculated based on the preprocessed BOLD: framewise displacement (FD), DVARS and three region-wise global signals extracted from CSF, WM, and whole-brain masks. FD was computed using two formulations following Power (absolute sum of relative motions) ^38^ and Jenkinson (relative root mean square displacement between affines) ^37^. FD and DVARS were calculated for each functional run, both using their implementations in Nipype. Frames that exceeded a threshold of 0.5 mm FD or 1.5 standardized DVARS were annotated as motion outliers. The head-motion estimates calculated in the correction step were also placed within the corresponding confounds file. The confound time series derived from head motion estimates and global signals were expanded with the inclusion of temporal derivatives and quadratic terms for each ^39^. All resampling was performed with a single interpolation step by composing all the pertinent transformations (i.e. head-motion transform matrices, susceptibility distortion correction when appropriate, and co-registrations to anatomical and output spaces). Gridded (volumetric) resampling was performed using antsApplyTransforms (ANTs), configured with Lanczos interpolation to minimize the smoothing effects of other kernels ^40^.

Because fMRIPrep does not apply temporal filtering, the same high-pass or bandpass filters as in the other pipelines were applied to the smoothed, MNI-space output prior to tSNR calculation. Confound time series were not regressed out at this stage.

#### 2.5.2. Preprocessing in CONN

The functional data was realigned using the SPM12 realign & unwarp procedure ^41^, which includes co-registration and resampling of all scans to a reference image (the first scan of the first session) through b-spline interpolation. This process also estimates the derivatives of the deformation field with respect to head movement, which allows addressing susceptibility associated with distortion-by-motion interactions. Moreover, the functional data is resampled to match the deformation field of the reference image. Finally, this step involves resampling the functional data along the phase-encoded direction, which corrects for the absolute deformation state of the reference image caused by field inhomogeneities in the MR scanner.

The CONN Toolbox identifies potential outlier volumes based on the observed global BOLD signal and the amount of subject-motion in the scanner. Acquisitions with framewise displacement (FD) above 0.9mm or global BOLD signal changes above 5 standard deviations are flagged as potential outliers (note: alternative "conservative" settings in CONN use 0.5mm and 3 standard deviation thresholds, while alternative "liberal" settings use 2mm and 9 standard deviation thresholds).

FD is computed at each timepoint by considering a 140x180x115mm bounding box around the brain and estimating the largest displacement among six control points placed at the center of the bounding-box faces. Global BOLD signal change is computed at each timepoint as the change in average BOLD signal within SPM’s global-mean mask scaled to standard deviation units.

The CONN Toolbox normalizes functional data into standard MNI space and segments them into gray matter, white matter, and CSF tissue classes using the SPM12 unified segmentation and normalization procedure ^42^. The procedure performs tissue classification iteratively, estimating the posterior tissue probability maps (TPMs) from the intensity values of the reference functional image and registration, estimating the nonlinear spatial transformation that best approximates the posterior and prior TPMs until convergence. The functional data is resampled to a default 180x216x180mm bounding box with 2mm isotropic voxels, using 4^th^-order spline interpolation.

Finally, the CONN Toolbox performs spatial smoothing on the functional data by convolving it with a Gaussian kernel of 6mm full width at half maximum (FWHM). Following this procedure, the same temporal filters, i.e. highpass and bandpass, as before were applied to the data.

#### 2.5.3. Preprocessing in FSL

The T1 images from all three sites were corrected for intensity inhomogeneities and skull-stripped using *fsl_anat*. Briefly, this pipeline ensures the correct orientation of the images and uses FAST ^33^ for bias correction and FNIRT ^43^ for brain extraction. B0 fieldmaps and magnitude images were collected to perform susceptibility distortion correction of the Stanford data (GE scanner). The magnitude image was skull-stripped using BET and the fieldmap was converted from Hz to rad/s prior to use. In addition, the fieldmap was prepared using 2D median filtering and despiking, and 3D Gaussian smoothing (sigma=1mm). Phase difference and magnitude images were collected to perform susceptibility distortion correction of the Toronto data (Siemens scanner). The magnitude image was skull-stripped using BET before entering this file with the phase difference file into *fsl_prepare_fieldmap*. As mentioned previously, susceptibility distortion correction was conducted online for data collected at Cincinnati site (Philips scanner). To allow for signal stabilization; the first 6 volumes of data was manually removed from the data collected at Stanford, these initial volumes were removed online for data collected at Toronto and Cincinnati. The functional data was then preprocessed using FEAT ^44^ with motion correction using MCFLIRT ^37^ combined with unwarping (when appropriate) using BBR and FUGUE, linear registration (FLIRT ^35,37^) to the bias-corrected brain-extracted T1 image (BBR), and MNI152_2mm standard brain (12 DOF), the same highpass and bandpass temporal filtering as before, and spatial smoothing. Preprocessed data was also non-linearly standardized to MNI using FNIRT and the transform matrices calculated by FEAT before calculating the tSNR.

### 2.6. Denoising

Once the optimal preprocessing pipeline was defined, i.e. FSL preprocessing including bandpass temporal filtering (Figure 2), subsequent analyses were conducted to compare the effect of commonly used denoising algorithms on the quality of the preprocessed data. Selected denoising techniques included aCompCor ^20^, FSL FIX ^21,22^, and ICA-AROMA ^23,45^. The same quality assessment as in the previous step was used to define the optimal denoising technique.

#### 2.6.1. Denoising with aCompCor

The application of the aCompCor noise-reduction technique was performed using CONN after preprocessing the data with the FSL pipeline as defined in step 1. It included a principal component analysis (PCA) and General Linear Model (GLM) analysis ^20^. The PCA was performed on the signal time series extracted from the white matter and CSF to create 5 nuisance regressors for each tissue (CONN default). These regressors, along with one regressor per outlier volume (volumes with framewise displacement above 0.9 mm or global BOLD signal change above 5 standard deviations; the number of outlier regressors therefore varied across participants) and 12 parameters (6 realignment parameters and their first temporal derivatives), were then used to create the design matrix for the GLM analysis. This technique is based on two assumptions ^20^: (1) The activation present in the white matter and CSF is only physiological noise and not signal of interest; (2) Noise in the gray matter is correlated to noise in the white matter and CSF and can be filtered out by regressing time series of noise in the white matter and CSF.

#### 2.6.2. Denoising with FSL FIX

##### Training data

The same procedure as described in Hoeppli et al. ^3^ was followed here. In short, three participants, including 2 resting-state series each, were randomly selected at each site to custom train a FIX classifier out of the 30 included in the sample. Using a custom-trained classifier optimizes its efficiency on one’s data ^21^. These participants underwent the same FSL preprocessing with bandpass filter as described above. A Probabilistic Independent Component Analysis (PICA) using MELODIC (Multivariate Exploratory Linear Optimized Decomposition into Independent Components) ^46^ was then performed to extract 25 components to classify. The number of components was a priori defined. Two experts (MEH & SPD) classified the components as noise, signal, or unknown following Salimi-Khorshidi et al.’s recommendations ^21^. Once consensus was achieved, the categorized ICs were used to train a FIX classifier. Simultaneously, a leave-one-out test was performed to assess the efficiency of the classifier to accurately distinguish noisy components from signal ones. Results of this test (Table 4) were used to define the distinction threshold used in the denoising of the test dataset.

**Table 4.**
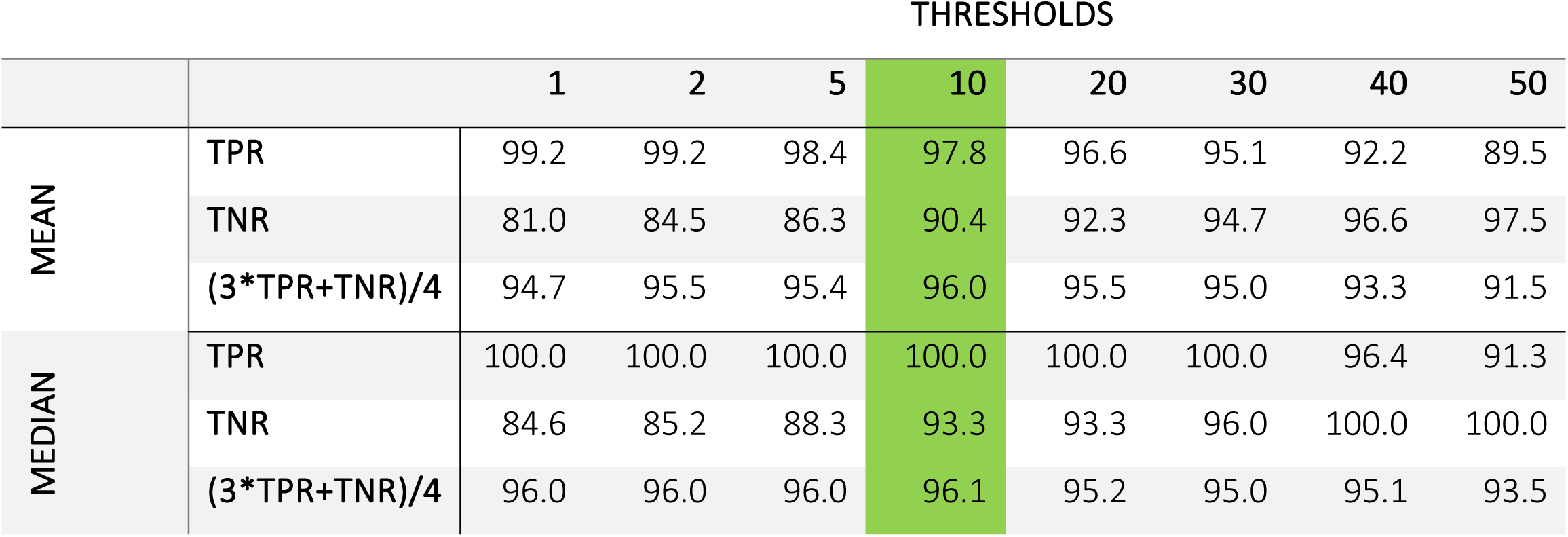
Results of the LOO test performed during the training of the FIX classifier. The selected distinction threshold for the denoising of the test dataset is highlighted in green. TPR: True Positive Rate; TNR: True Negative Rate.

##### Test data

After preprocessing with FSL including bandpass filtering, PICA using MELODIC was performed on the data to define 25 independent components in each of the remaining 21 participants. The custom-trained FIX classifier was then used to automatically classify and filter out the components defined by MELODIC into noise or signal with a distinction threshold of 10.

#### 2.6.3. Denoising with ICA-AROMA

ICA-AROMA was implemented using the Python 3 script developed by Pruim et al. ^23,45^. Although similar to FIX, ICA-AROMA does not require a custom-trained classifier. It identifies and filters out independent components related to motion and high-frequency fluctuations that are unlikely to represent neural activity.

### 2.7. Statistical analyses

A two-stage analytical approach was employed. In the first stage, preprocessing pipelines were compared using tSNR values extracted from the 10 ROIs. In the second stage, denoising methods were compared on the preprocessing pipeline selected in step 1. Each stage used the same statistical approach.

The primary statistical analysis used linear mixed-effects (LME) models to account for the repeated-measures structure of the data, where multiple ROI-level tSNR observations were nested within each participant. The LME model was specified as tSNR ∼ pipeline + ROI + (1|subject), with a random intercept per subject to capture between-subject variability. Pairwise comparisons between pipelines (or denoising methods) were obtained by refitting the model for each pair, with Bonferroni correction applied across the three pairwise tests.

To evaluate site-specific performance, a site × pipeline interaction term was added to the LME (tSNR ∼ pipeline * site + ROI + (1|subject)), and the significance of the interaction was assessed by comparing models with and without the interaction term using likelihood ratio tests (LRT). Cross-site variability per pipeline was quantified as the coefficient of variation (CV) of the site-level mean tSNR values.

As a complementary analysis, the subject-level mean tSNR (averaged across ROIs) was also examined. Normality was assessed per group using the Shapiro-Wilk test (alpha = 0.05). When all groups satisfied the normality assumption, a one-way ANOVA served as the omnibus test; when any group violated normality, the Kruskal-Wallis H test was used. Variance homogeneity was assessed using the Brown-Forsythe test. The filter effect (bandpass filter versus high-pass filter) was tested within each pipeline. Differences in tSNR and motion parameter (FD) across sites were tested using the Kruskal-Wallis H test. Two outliers identified via the 1.5 times interquartile range rule were excluded from denoising visualizations but retained in the statistical tests, which were computed on the full dataset. Analyses were performed in Python 3.14.7 and figures were generated in R 4.6.1.

Analysis scripts and statistical maps produced for this manuscript are publicly available at https://osf.io/nufw5/overview. Defaced raw image data will be made publicly available as part of the SPRINT dataset release, in accordance with the SPRINT open data policy. The data repository and access details will be posted on the same OSF page once the release is complete.

## 3. Results

### 3.1. Acquisition sites differ in tSNR and head motion

Before evaluating preprocessing pipelines, we characterized signal quality and head motion across the three acquisition sites (Figure 3). Sites differed significantly in mean tSNR after FSL-based preprocessing with bandpass filter (Kruskal-Wallis H = 6.80, p = 0.033), with Cincinnati showing the highest values (M = 482.0, SD = 162.9), followed by Toronto (M = 339.2, SD = 103.3) and Stanford (M = 293.5, SD = 129.6). Sites also differed significantly in head motion (H = 7.38, p = 0.025), with Cincinnati exhibiting the highest mean framewise displacement (M = 0.27, SD = 0.09 mm) and Stanford the lowest (M = 0.15, SD = 0.12 mm; Toronto: M = 0.17, SD = 0.07 mm). Notably, Cincinnati showed simultaneously higher tSNR and higher motion. If head motion were the primary driver of site-level tSNR differences, the expected direction would be reversed; this pattern instead suggests that scanner- or acquisition-related factors underlie the observed site-level variation in signal quality.

**Figure 3:**
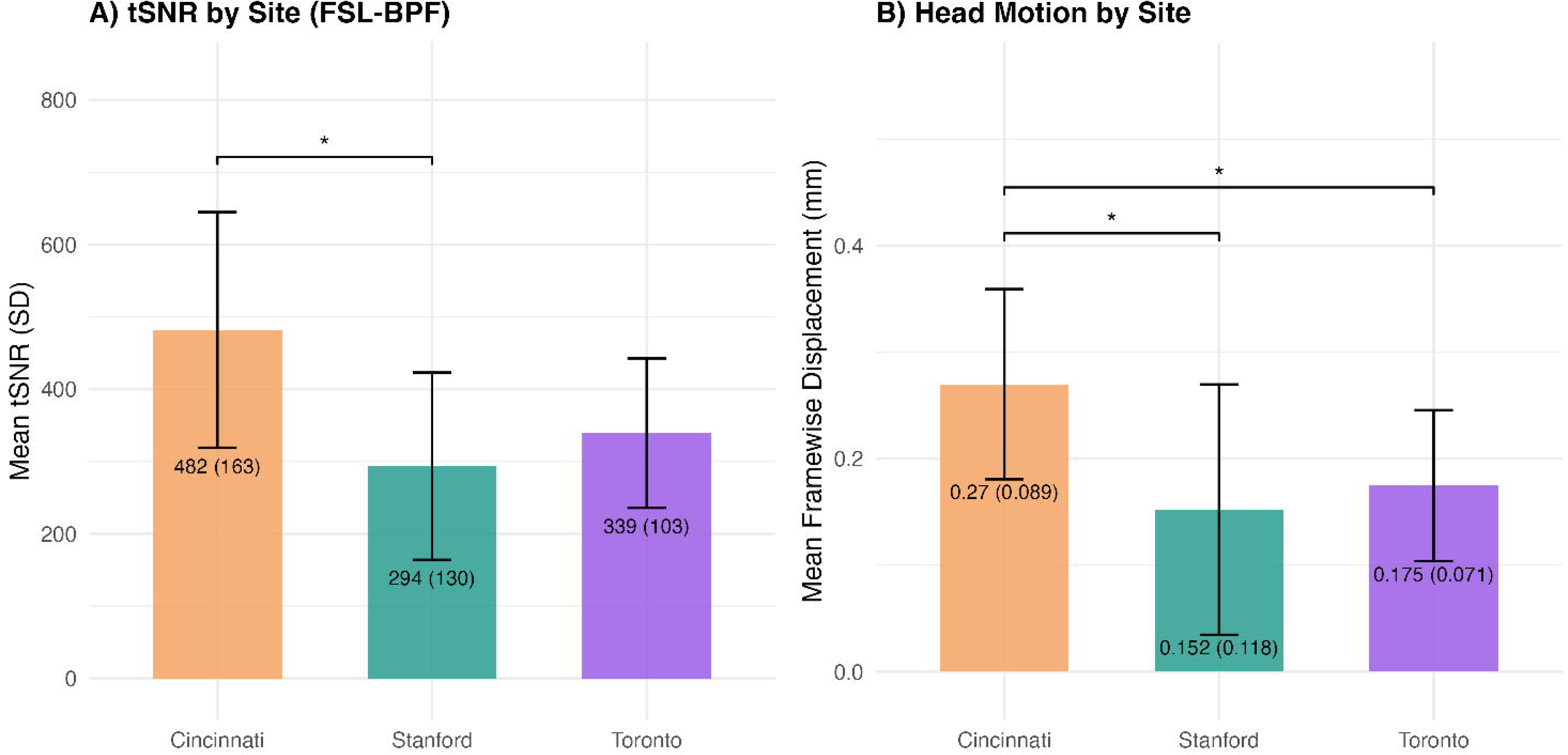
Site characterization. (A) Mean tSNR under FSL-bandpass filter preprocessing by acquisition site (mean ± SD). Sites differed significantly in tSNR (Kruskal-Wallis H = 6.80, p = 0.033); the bracket indicates a Bonferroni-corrected significant difference between Cincinnati and Stanford (p = 0.046). (B) Mean framewise displacement (FD) by acquisition site (mean ± SD). Sites differed significantly in head motion (Kruskal-Wallis H = 7.38, p = 0.025); brackets indicate Cincinnati was significantly higher than both Stanford and Toronto (Bonferroni-corrected p values). The co-occurrence of higher tSNR and higher motion in Cincinnati suggests that scanner- or acquisition-related factors, rather than participant motion, underlie the site-level differences in signal quality.

### 3.2. Bandpass filtering yields higher tSNR than high-pass filtering across all pipelines

We evaluated whether <u>bandpass filter</u> (0.008 Hz – 0.09 Hz) or high-pass filter produced higher tSNR, as this choice affects all downstream comparisons. Bandpass filtering produced significantly higher tSNR than high-pass filtering across all three pipelines (Figure 4). The effect was most pronounced for FSL, which showed a 2.26-fold increase with bandpass filter relative to HPF (i.e., bandpass filter tSNR was 2.26 times the high-pass filter value; p < 0.001), compared to 1.43-fold for fMRIPrep (p = 0.006) and 1.42-fold for CONN (p = 0.001). These consistent and significant improvements indicate that removing high-frequency fluctuations through bandpass filtering substantially increases temporal signal stability regardless of pipeline. All subsequent pipeline and denoising comparisons therefore used bandpass-filtered data.

**Figure 4:**
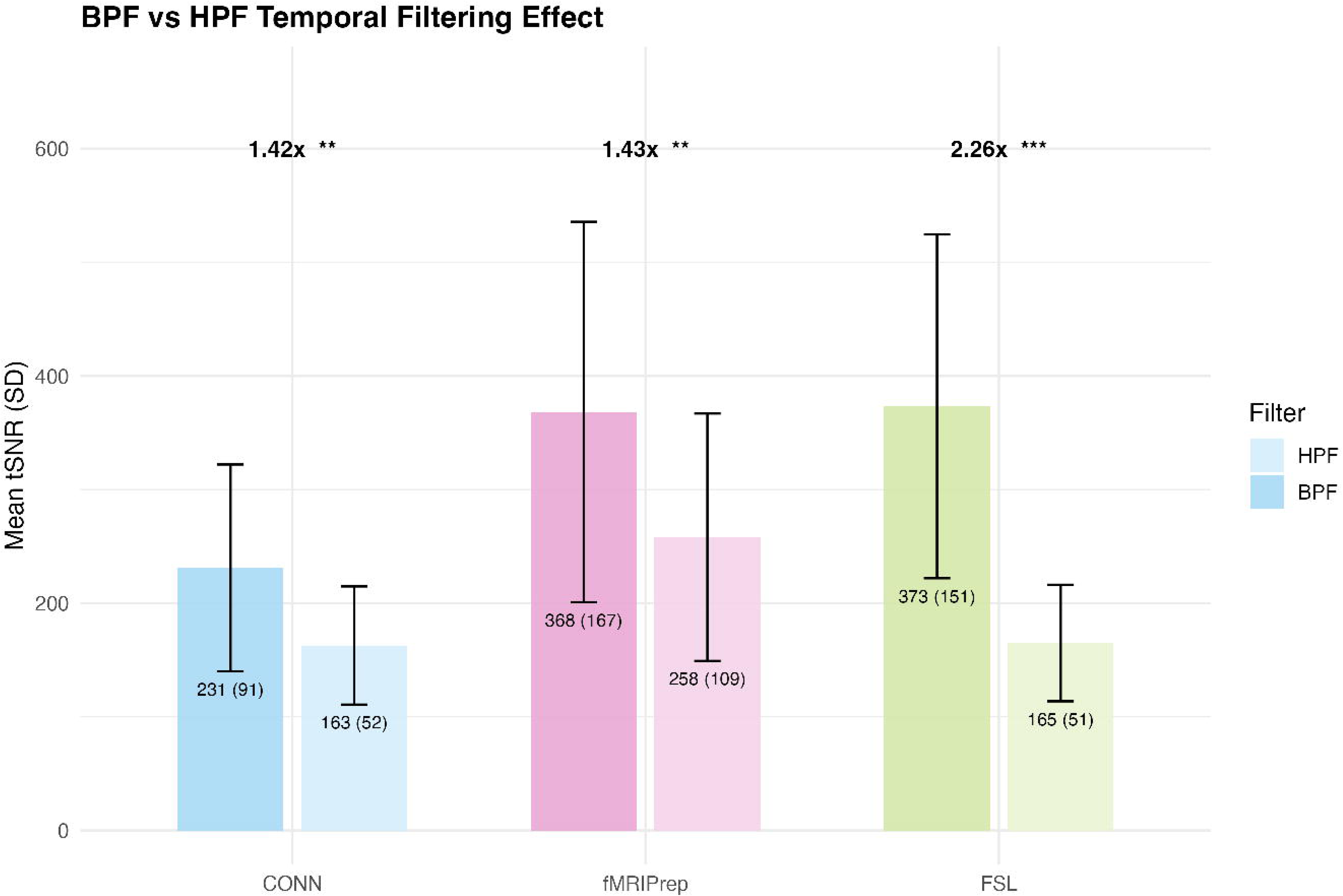
Temporal filtering effect (bandpass filter vs high-pass filter). Mean tSNR under bandpass filter (BPF, darker bars) and high-pass filter (HPF, lighter bars) filtering for each preprocessing pipeline (CONN, fMRIPrep, FSL), with error bars indicating standard deviation. Fold-change and significance of the bandpass filter vs high-pass filter comparison are shown above each pipeline pair (Bonferroni-corrected paired comparisons; ** p < 0.01, *** p < 0.001). Bandpass filter consistently outperformed high-pass filter across all three pipelines, with the largest gain in FSL (2.26-fold).

### 3.3. FSL preprocessing pipeline presented higher tSNR with superior consistency

Having established that bandpass filter outperforms HPF, we compared the effect of the three preprocessing pipelines on bandpass-filtered data. The LME model revealed significant effects of the preprocessing pipeline on regional tSNR when bandpass filtering was applied (10 ROIs, 810 observations; Figure 5C). Bonferroni-corrected pairwise LME comparisons indicated that both FSL (coefficient = 142.2, p < 0.001) and fMRIPrep (coefficient = 137.2, p < 0.001) produced significantly higher tSNR than CONN, while FSL and fMRIPrep did not differ significantly (coefficient = -5.0, p = 1.0). Descriptively, FSL showed the highest mean tSNR (M = 373.3, SD = 151.3), followed by fMRIPrep (M = 368.2, SD = 167.4) and CONN (M = 231.0, SD = 91.0). The complementary subject-mean ANOVA corroborated these findings (F(2,78) = 8.91, p < 0.001). The tSNR distributions satisfied the normality assumption (Shapiro-Wilk, all p > 0.12; Figure 5A), with a trend toward heterogeneous variances (Brown-Forsythe F = 2.68, p = 0.075). This pattern was consistent across the regional tSNR profile (Figure 5B), where FSL and fMRIPrep showed higher tSNR than CONN in all 10 ROIs. Brain surface visualization confirmed these differences, with CONN showing uniformly lower cortical tSNR compared to fMRIPrep and FSL (Figure 5D).

**Figure 5:**
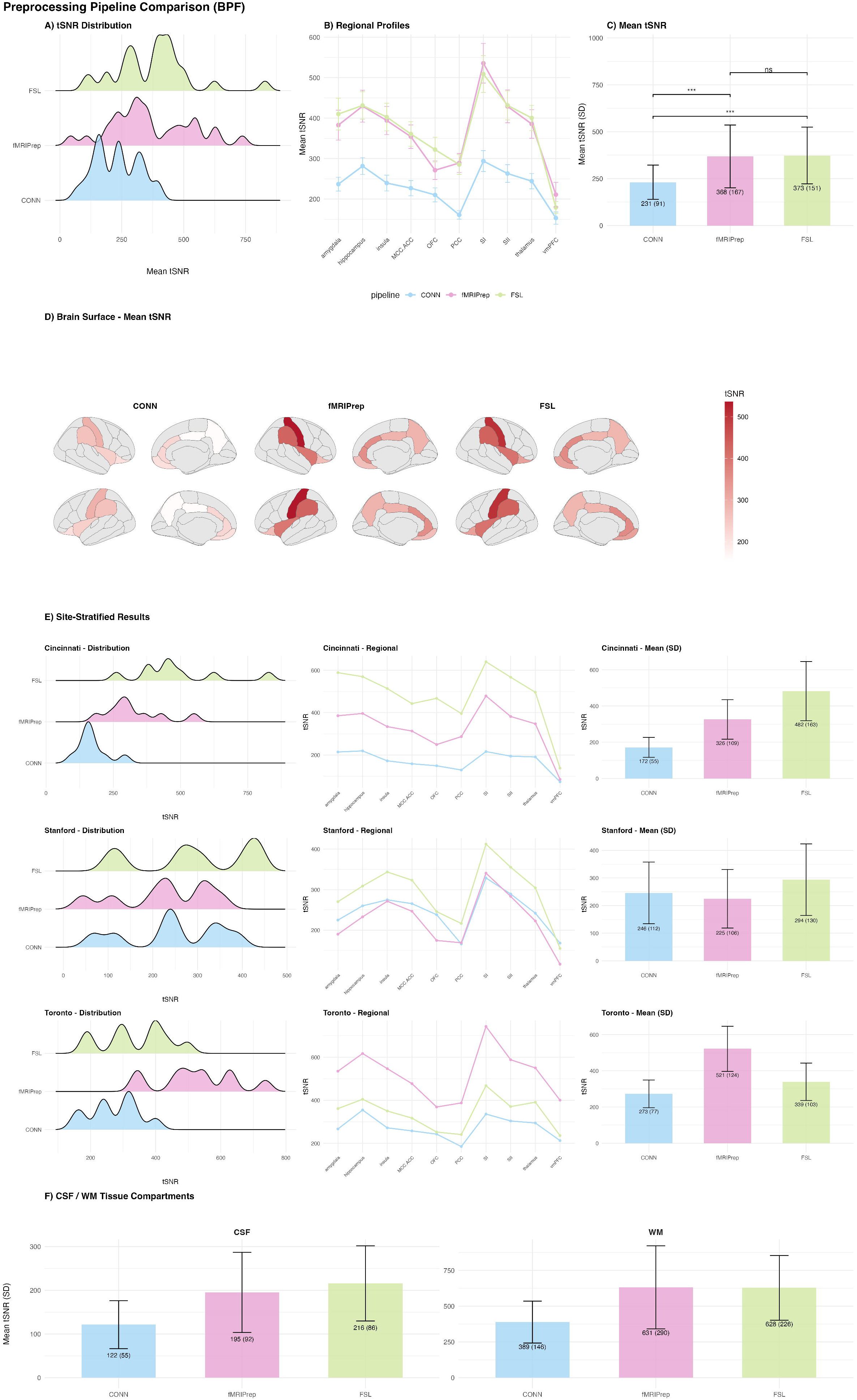
Comparison of the 3 preprocessing pipelines (FSL, fMRIPrep, CONN). (A) Density ridge plots showing the distribution of mean tSNR across the 10 pain-relevant ROIs for each preprocessing pipeline (FSL, fMRIPrep, CONN) under bandpass filtering. (B) Regional tSNR profiles across the 10 ROIs for each pipeline under bandpass filter, showing mean and standard error. (C) Mean tSNR by pipeline under bandpass filter, with error bars indicating standard deviation and significance brackets from Bonferroni-corrected t-tests (** p < 0.01, *** p < 0.001, ns = not significant). (D) Cortical brain surface maps (Desikan-Killiany atlas) displaying mean tSNR for each pipeline, with a shared white-to-red color scale. (E) Site-stratified results showing tSNR distributions, regional profiles, and mean values separately for Cincinnati, Stanford, and Toronto. (F) Mean tSNR in cerebrospinal fluid (CSF) and white matter (WM) tissue compartments by pipeline.

Tissue compartment analysis (Figure 5F) showed that FSL and fMRIPrep both achieved higher tSNR in white matter (FSL: M = 628, SD = 226; fMRIPrep: M = 631, SD = 290) and CSF (FSL: M = 216, SD = 86; fMRIPrep: M = 195, SD = 92) compared to CONN (WM: M = 389, SD = 146; CSF: M = 122, SD = 55). The replication of this pattern in non-neural tissue indicates that the tSNR advantage of FSL and fMRIPrep reflects global noise suppression properties of the pipelines, rather than being specific to the cortical and subcortical ROIs examined. This is of relevance because white matter and CSF signals serve as noise references in subsequent denoising strategies such as aCompCor; more temporally stable reference signals directly improve the quality of nuisance regression.

### 3.4. Optimal pipeline differs by site: FSL leads in Cincinnati and Stanford, fMRIPrep in Toronto

A critical finding of this study is that the optimal preprocessing pipeline differed across acquisition sites (Figure 6). A likelihood ratio test revealed a highly significant site × pipeline interaction across all three pipelines (chi²(4) = 526.31, p < 0.001). Examining site-level performance separately, FSL yielded the highest tSNR in Cincinnati (M = 482.0, SD = 163.0; ANOVA F(2,24) = 15.65, p < 0.001), whereas in Stanford the three pipelines did not differ significantly (F(2,21) = 0.74, p = 0.49) with FSL showing the highest mean (M = 293.5, SD = 129.6). In contrast, fMRIPrep substantially outperformed FSL in Toronto (M = 521.3, SD = 124.3 vs. FSL M = 339.2, SD = 103.3; ANOVA F(2,27) = 15.53, p < 0.001). This crossover interaction indicates that a pipeline’s performance cannot be assumed to generalize across scanners, and that single-pipeline recommendations should be validated per site in multisite designs with heterogeneous hardware.

**Figure 6:**
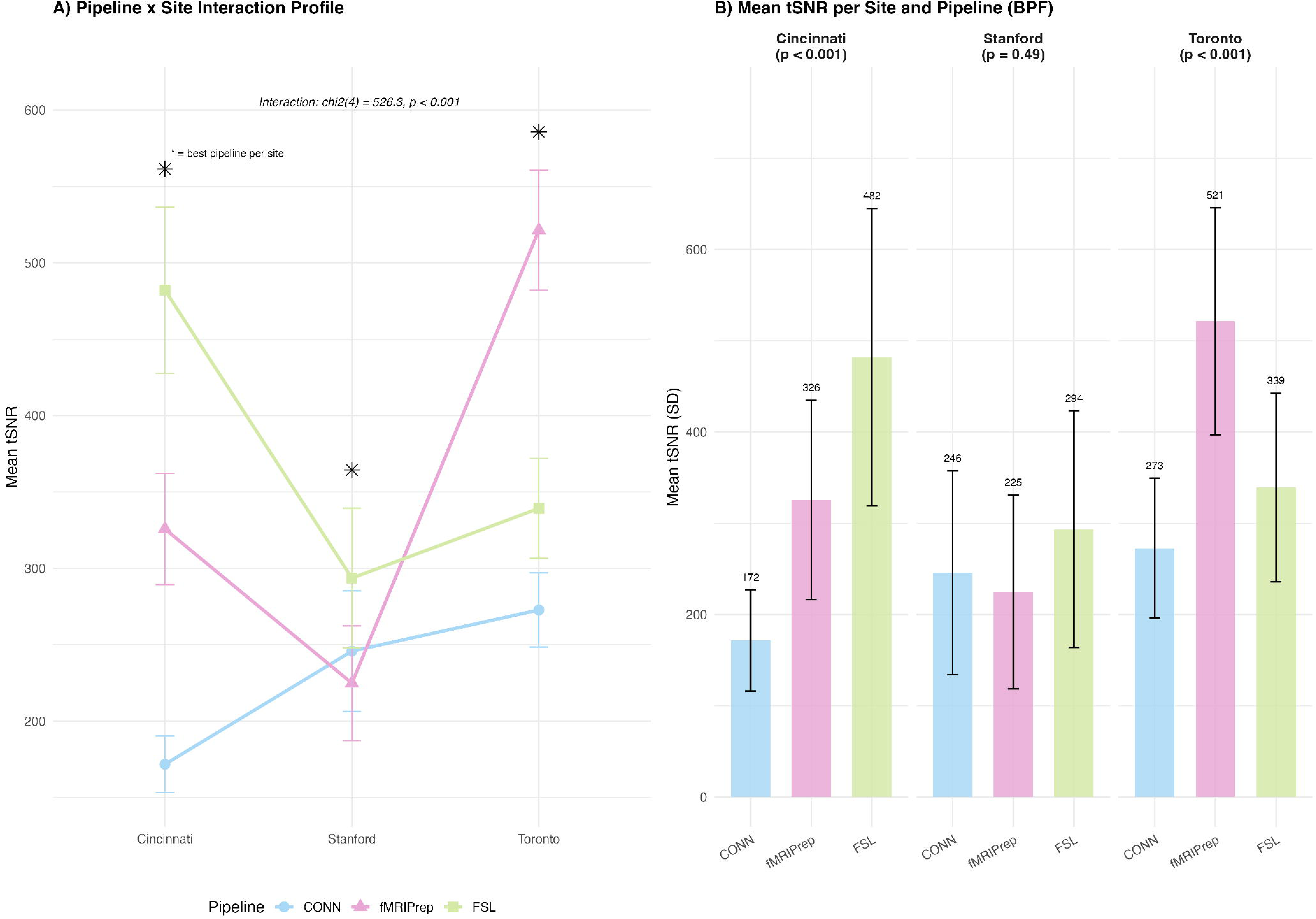
Site × pipeline interaction. (A) Interaction profile showing mean tSNR per site for each preprocessing pipeline (lines connect site means; * indicates the best-performing pipeline per site; chi²(4) = 526.31, p < 0.001). (B) Grouped bar charts per site showing mean tSNR for all three pipelines under bandpass filter, with omnibus ANOVA p-values per site. FSL performed best in Cincinnati and Stanford; fMRIPrep performed best in Toronto, highlighting the dependence of optimal pipeline choice on scanner-or acquisition-related characteristics.

The degree of cross-site variability also differed markedly between pipelines. fMRIPrep exhibited the largest cross-site variability (CV = 34.5%, site mean range = 296.6), suggesting strong interactions with scanner-specific characteristics. FSL showed intermediate variability (CV = 21.6%, range = 188.5), while CONN —despite having the lowest overall tSNR— was the most consistent across sites (CV = 18.6%, range = 101.0). The pairwise LRT comparing FSL and fMRIPrep alone confirmed a significant interaction (chi²(2) = 327.88, p < 0.001).

Based on these combined results, FSL with bandpass filtering was selected as the preprocessing pipeline for the denoising comparison, given its superior overall tSNR relative to CONN, its strongest bandpass filter response, and its more consistent cross-site performance compared to fMRIPrep. This choice reflects the trade-off outlined in the Introduction: FSL was not the best pipeline at every site, but was the pipeline whose performance was least dependent on site.

### 3.5. FIX achieves higher tSNR than ICA-AROMA and aCompCor

The LME model revealed significant differences between denoising methods when accounting for within-subject ROI-level variability (10 ROIs, 750 observations; Figure 7C). Bonferroni-corrected pairwise LME comparisons showed that FIX produced significantly higher tSNR than both ICA-AROMA (coefficient = 57.7, p < 0.001) and aCompCor (coefficient = 74.4, p < 0.001), while ICA-AROMA and aCompCor did not differ significantly (coefficient = 16.7, p = 0.38). Descriptively, FIX showed the highest tSNR (M = 396.2, SD = 164.5), followed by ICA-AROMA (M = 338.5, SD = 114.6) and aCompCor (M = 321.8, SD = 131.9). The complementary subject-mean Kruskal-Wallis test did not reach significance (H = 3.91, p = 0.14), reflecting the lower statistical power of collapsing ROI-level data into a single mean per subject. The Shapiro-Wilk test indicated non-normal distributions for FIX (W = 0.78, p < 0.001) and aCompCor (W = 0.89, p = 0.014), while variances were homogeneous (Brown-Forsythe F = 0.06, p = 0.94).

**Figure 7:**
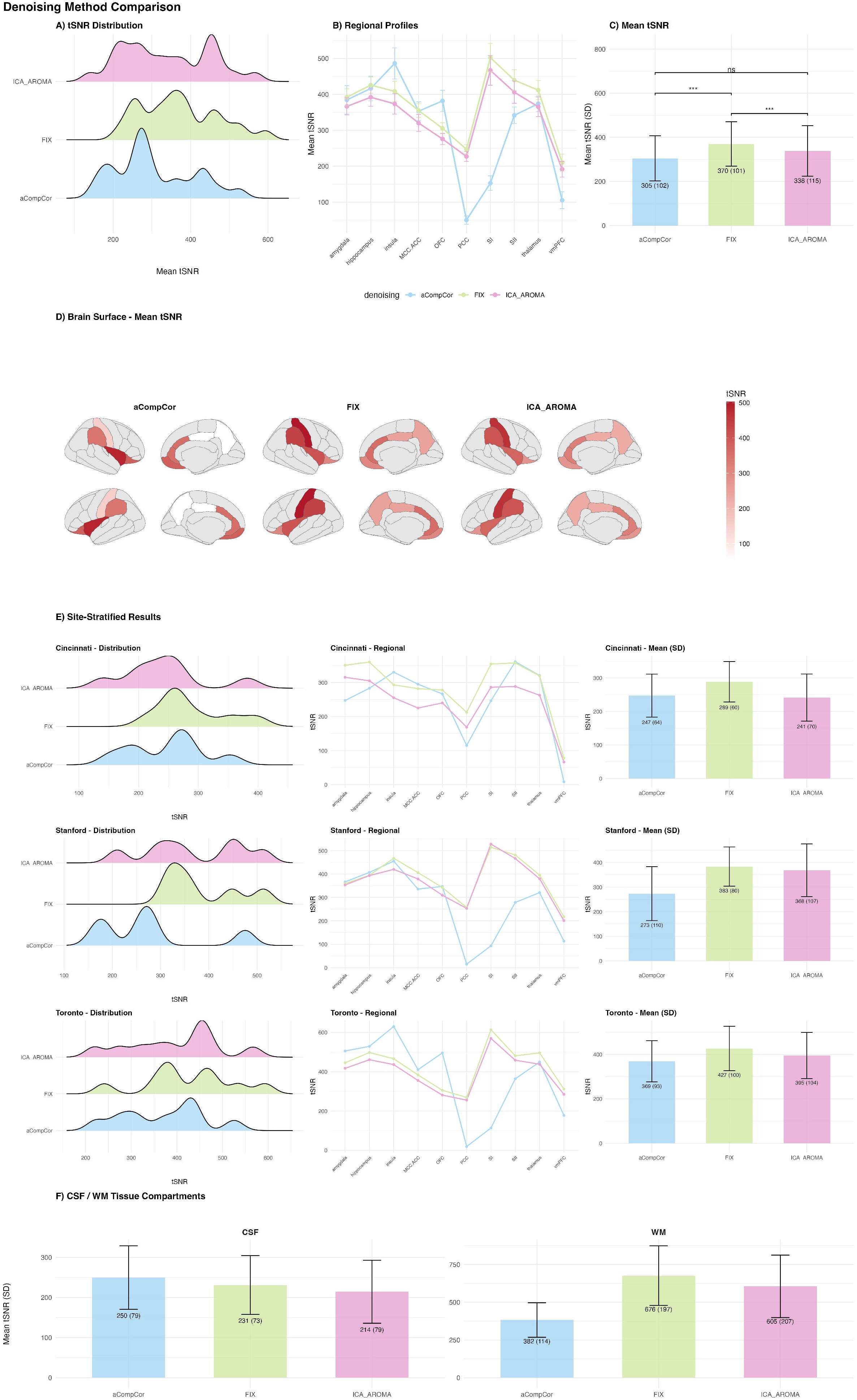
Comparison of the 3 denoising methods (aCompCor, FIX, ICA-AROMA). (A) Density ridge plots showing the distribution of mean tSNR for each denoising method (aCompCor, FIX, ICA-AROMA) applied to FSL-preprocessed data with bandpass filtering. Two statistical outliers were removed from visualizations. (B) Regional tSNR profiles across the 10 ROIs for each denoising method, showing mean and standard error. (C) Mean tSNR by denoising method, with error bars indicating standard deviation and significance brackets from Bonferroni-corrected Mann-Whitney U tests (ns = not significant). Statistical tests were performed on the full dataset. (D) Cortical brain surface maps (Desikan-Killiany atlas) displaying mean tSNR for each denoising method, with a shared white-to-red color scale. (E) Site-stratified results showing tSNR distributions, regional profiles, and mean values separately for Cincinnati, Stanford, and Toronto. (F) Mean tSNR in CSF and WM tissue compartments within denoising method.

The regional tSNR profile (Figure 7B) showed a consistent pattern across methods, with FIX producing higher values than ICA-AROMA across all 10 ROIs (difference range: 34–81 tSNR units), with the largest advantages in primary somatosensory cortex (SI; FIX: M = 548, ICA-AROMA: M = 467), mid-cingulate/anterior cingulate cortex (MCC-ACC; FIX: M = 388, ICA-AROMA: M = 320), and thalamus (FIX: M = 430, ICA-AROMA: M = 366). The advantage of FIX over aCompCor was more variable across regions: FIX showed substantially higher tSNR in SI (M = 548 vs. M = 161), PCC (M = 282 vs. M = 49), and SII (M = 467 vs. M = 376), while aCompCor exceeded FIX in insula (M = 508 vs. M = 436) and OFC (M = 396 vs. M = 327), regions anatomically proximal to CSF compartments that aCompCor explicitly models. Brain surface visualization (Figure 7D) confirmed the overall pattern, with FIX showing higher cortical tSNR compared to ICA-AROMA and aCompCor across the majority of the cortex.

Site-stratified analyses (Figure 7E) did not reveal significant differences between denoising methods within any individual site (Cincinnati: F(2,24) = 1.25, p = 0.31; Stanford: H = 3.44, p = 0.18; Toronto: F(2,27) = 0.84, p = 0.44).

Tissue compartment analysis (Figure 7F) indicated that aCompCor yielded the highest CSF tSNR (M = 267, SD = 117) compared to FIX (M = 245, SD = 100) and ICA-AROMA (M = 214, SD = 79), while FIX showed the highest WM tSNR (M = 702, SD =231). The relatively higher CSF tSNR under aCompCor is expected given that this method explicitly models CSF signal components in its regression framework.

In summary, the LME analysis identified FIX as the denoising method producing significantly higher tSNR compared to both ICA-AROMA and aCompCor when applied to FSL-preprocessed data with bandpass filtering. The advantage of the LME approach over the subject-mean analysis is that it leverages the full ROI-level data structure while properly accounting for within-subject correlation, providing greater statistical power without pseudo-replication.

## 4. Discussion

In this study, we aimed to define the optimal preprocessing pipeline for a large multisite dataset. Data was acquired at three different sites, i.e. the Richard M. Lucas Center for Imaging at Stanford School of Medicine (GE scanner), Cincinnati Children’s Hospital Medical Center (Philips scanner), and SickKids Hospital Toronto (Siemens scanner). Each site owned a scanner from a different brand, which implies differences in the acquisition parameters, despite our efforts to program similar scanning sequences, and in the acquired data as a result. Although we aimed at minimizing these differences during the definition of the sequences and data acquisition, we needed to carefully consider the preprocessing pipeline to achieve two preprocessing goals: (1) ensure that data from each site was preprocessed optimally, and (2) harmonize the preprocessing pipeline across sites, so that all data are preprocessed similarly. A balance needed to be struck between these two goals (site-specific preprocessing and harmonization). This is unlike previous studies, which primarily focused on harmonization of very specific preprocessing steps between sites, in particular for intensity normalization ^2,11^. Our analyses showed significant differences in noise level between sites in the raw data, highlighting Cincinnati as the site with the highest tSNR and the greatest head motion. This suggests a non-negligible role of scanner or sequence parameters in the observed noise. Based on this finding, it was essential to ensure the optimization of the preprocessing pipeline for each site. Overall, our results showed that an FSL-based preprocessing pipeline, followed by a denoising using a FIX custom-trained classifier achieved optimal preprocessing of the data at each site, while harmonizing the pipeline across sites.

### 4.1 Preprocessing pipeline: effect of site

Our first step was to define the optimal preprocessing pipeline. For this purpose, we compared three commonly used pipelines: fMRIPrep, CONN, and FSL, and investigated the effect of the type of temporal filtering on the efficiency of the preprocessing, i.e. bandpass vs. high-pass filtering. Results of this comparison showed a clear advantage of applying bandpass filtering over high-pass filtering in all three preprocessing pipelines, i.e. FSL, fMRIPrep, and CONN: data preprocessed with bandpass filter had significantly greater mean tSNR values than data preprocessed using high-pass filter. That effect was the greatest in the FSL pipeline. Bandpass filtering has traditionally been used in the preprocessing of resting-state fMRI data. However, a recent study suggested that the application of a high-pass filter yielded similar results and allowed to conserve signal of interest that might be present at higher frequencies ^47^. Our results differ from this this prior finding, although the two studies are not directly comparable: Shirer et al.^47^ evaluated network detection and test-retest reliability after nuisance regression that included individual physiological recordings, whereas we evaluated tSNR on data without nuisance regression. In that setting, the high-frequency variance that a bandpass filter removes is precisely what physiological regressors, as the ones used in Shirer et al., would otherwise account for. This likely explains the larger benefit of bandpass filtering in our data. It should also be kept in mind that tSNR increases mechanically when temporal variance is reduced, so part of this gain reflects the metric rather than improved signal. Based on our results, all the following analyses were performed with a bandpass filter.

The comparison of the three preprocessing pipelines highlighted 3 findings. First, the mean tSNR following the FSL and fMRIPrep pipelines was greater than after CONN, with FSL slightly outperforming fMRIPrep. Second, within-site analyses showed that FSL outperformed the other pipelines in Stanford and Cincinnati, while fMRIPrep outperformed the others in Toronto. Third, FSL and CONN had less cross-site variability than fMRIPrep.

These results highlight some of the characteristics for which the FSL and fMRIPrep pipelines are well known. FSL is typically considered a very flexible pipeline that allows to adjust the fit of the pipeline precisely to one’s data. In contrast, fMRIPrep is recognized as a standardized pipeline that can support better reproducibility of scientific findings ^48^. Because of this, fMRIPrep is often chosen as the go-to preprocessing pipeline. Our results suggest that the need to balance harmonization of the preprocessing with site-specific needs in multisite datasets might be better met by a more flexible approach, such as the one provided by FSL. Therefore, researchers preprocessing multisite data should carefully consider the choice of the preprocessing pipeline and thoroughly investigate its outcome to ensure optimal preprocessing of their data.

Based on all these results, FSL with bandpass filter was chosen as the optimal preprocessing pipeline for our data, which allowed efficient removal of the noise and reduced cross-site variability. This pipeline was used as the basis for the comparison of the denoising techniques.

### 4.2 Denoising

In the second step, we compared three denoising techniques applied to the data preprocessed with the the FSL pipeline as defined above: aCompCor, FSL FIX, and ICA-AROMA. Results from this step highlighted the overall significant benefit of denoising with FSL FIX over the other denoising techniques. Interestingly, FIX performed better within and across sites. Our results add to previous findings ^3^. Indeed, Hoeppli et al. demonstrated the benefit of using FSL FIX to achieve a good balance between removing noise and conserving the signal of interest in an fMRI dataset collected at a single site. Our current results extend this knowledge by demonstrating the value of using FSL FIX to achieve such a balance in a multisite setting. This demonstration is especially important given the current increase in multisite studies including the acquisition of fMRI data.

In addition to its greater efficiency in denoising the data, the FIX classifier is a technique that is sensitive to the specific spatial and temporal features of one’s dataset, especially when the classifier is custom-trained. This sensitivity is particularly important when working on resting-state fMRI sequences. Resting-state fMRI data has been previously associated with limited reliability ^12^. In their study, Schwartz et al. highlighted a lack of reliability within and between sites, even though all sites in the study had a Siemens scanner. These findings further emphasize the need to carefully confirm the efficiency and sensitivity of the chosen preprocessing pipeline and denoising technique.

In our study setting, the use of FIX after the FSL preprocessing pipeline has two additional advantages. First, the definition of a custom-trained classifier using data from all sites allows for a more harmonized denoising of the data across sites. Second, the defined preprocessing pipeline and the denoising technique are both FSL-based, which limits the risk of incompatibility between tools.

To our knowledge, this is the first study aimed at comparing preprocessing pipelines, including denoising techniques, in their entirety for multisite fMRI data. In short, an FSL-based preprocessing pipeline followed by denoising with a custom-trained FSL FIX classifier allowed us to achieve an optimal balance between harmonization of the process between sites and site-specific needs. Although our results can be a guide for other researchers, a careful *a priori* investigation of the effect of the chosen preprocessing pipeline on their data is recommended.

One limitation of our study is the lack of testing of individual preprocessing steps using different tools. Our data did not require such customization of the preprocessing pipelines. Therefore, aside from the temporal filtering, we focused on the preprocessing pipeline in its entirety. However, other researchers might benefit from such flexibility in the definition of an optimal preprocessing pipeline for their datasets. Furthermore, while tSNR is a robust metric for evaluating foundational data quality and noise reduction, we acknowledge that higher tSNR does not intrinsically guarantee improved detection of resting-state networks. Extremely aggressive filtering can sometimes inflate tSNR while inadvertently removing neurobiologically relevant signals. Future steps in this multisite project might evaluate how these optimized preprocessing and denoising pipelines directly impact functional connectivity matrices and the identifiability of specific resting-state networks.

## 5. Conclusions

To the best of our knowledge, our study is the first to define a preprocessing pipeline for multisite resting-state fMRI data, which aims at balancing harmonization across sites with site-specific needs. Our results show that the best performing pipeline can differ between scanners, and that an FSL-based preprocessing pipeline followed by denoising using a custom-trained FIX classifier achieved the best balance between data quality and cross-site consistency in our dataset. These results can support other researchers in the definition of an optimal pipeline for their datasets, with the recommendation that candidate pipelines be tested per site before one is adopted.

## Acknowledgments

This work was funded by the National Center for Complementary and Integrative Health, U.S. National Institutes of Health, under awards R61NS114926 and R33NS114926-02. Saül Pascual-Diaz received additional funding by the Spanish Ministry of Science and Innovation (MCIN/AEI, 10.13039/501100011033) through grants PID2020-117327RB-I00, PID2023-147704OB-I00, and CNS2023- 145425.

This work would not have been possible without the hard work of research coordinators who completed all the data collection.

## References

1. Zhou HH, Singh V, Johnson SC, et al. Statistical tests and identifiability conditions for pooling and analyzing multisite datasets. Proc Natl Acad Sci 2018;115:1481–6.

2. Wachinger C, Rieckmann A, Pölsterl S, ageing for the ADNI and the AIB and L flagship study of. Detect and correct bias in multi-site neuroimaging datasets. Méd Image Anal 2021;67:101879.

3. Hoeppli ME, Garenfeld MA, Mortensen CK, Nahman-Averbuch H, King CD, Coghill RC. Denoising task-related fMRI: Balancing noise reduction against signal loss: data repository. Human Brain Mapping 2023:1–1.

4. Moayedi M, Salomons TV, Atlas LY. Pain Neuroimaging in Humans: a Primer for Beginners and Non-Imagers. Journal of Pain March 2018:1–62.

5. Hoeppli ME, Garenfeld MA, Mortensen CK, Nahman-Averbuch H, King CD, Coghill RC. Denoising task-related fMRI: Balancing noise reduction against signal loss. Hum Brain Mapp 2023.

6. Moses P, Hernandez LM, Orient E. Age-related differences in cerebral blood flow underlie the BOLD fMRI signal in childhood. Front Psychol 2014;5:300.

7. Tian D, Zeng Z, Sun X, et al. A deep learning-based multisite neuroimage harmonization framework established with a traveling-subject dataset. NeuroImage 2022;257:119297.

8. Bento M, Fantini I, Park J, Rittner L, Frayne R. Deep Learning in Large and Multi-Site Structural Brain MR Imaging Datasets. Front Neuroinformatics 2022;15:805669.

9. Larivière S, Paquola C, Park B, et al. The ENIGMA Toolbox: multiscale neural contextualization of multisite neuroimaging datasets. Nat Methods 2021;18:698–700.

10. Wrobel J, Martin ML, Bakshi R, et al. Intensity warping for multisite MRI harmonization. NeuroImage 2020;223:117242.

11. Torbati ME, Minhas DS, Ahmad G, et al. A multi-scanner neuroimaging data harmonization using RAVEL and ComBat. NeuroImage 2021;245:118703.

12. Schwartz DL, Tagge I, Powers K, et al. Multisite reliability and repeatability of an advanced brain MRI protocol. J Magn Reson Imaging 2019;50:878–88.

13. Simons L, Moayedi M, Coghill RC, et al. Signature for Pain Recovery IN Teens (SPRINT): Protocol for a multi-site prospective signature study in chronic musculoskeletal pain. BMJ 2022.

14. Esteban O, Markiewicz CJ, Blair RW, et al. FMRIPrep: a robust preprocessing pipeline for functional MRI. Nat Methods 2019;16:111–6.

15. Esteban O, Ciric R, Finc K, et al. Analysis of task-based functional MRI data preprocessed with fMRIPrep. Nat Protoc 2020;15:2186–202.

16. Woolrich MW, Jbabdi S, Patenaude B, et al. Bayesian analysis of neuroimaging data in FSL. NeuroImage 2009;45:S173–86.

17. Smith SM, Jenkinson M, Woolrich MW, et al. Advances in functional and structural MR image analysis and implementation as FSL. NeuroImage 2004;23:S208–19.

18. Jenkinson M, Beckmann CF, Behrens TEJ, Woolrich MW, Smith SM. FSL. NeuroImage 2012;62:782–90.

19. Nieto-Castanon A. FMRI minimal preprocessing pipeline. In: Handbook of Functional Connectivity Magnetic Resonance Imaging Methods in CONN. Vol Hilbert Press; 2020:3–16.

20. Behzadi Y, Restom K, Liau J, Liu TT. A component based noise correction method (CompCor) for BOLD and perfusion based fMRI. NeuroImage 2007;37:90–101.

21. Salimi-Khorshidi G, Douaud G, Beckmann CF, Glasser MF, Griffanti L, Smith SM. Automatic denoising of functional MRI data: combining independent component analysis and hierarchical fusion of classifiers. NeuroImage 2014;90:449–68.

22. Griffanti L, Salimi-Khorshidi G, Beckmann CF, et al. ICA-based artefact removal and accelerated fMRI acquisition for improved resting state network imaging. NeuroImage 2014;95:232–47.

23. Pruim RHR, Mennes M, van Rooij D, Llera A, Buitelaar JK, Beckmann CF. ICA-AROMA: A robust ICA-based strategy for removing motion artifacts from fMRI data. NeuroImage 2015;112:267–77.

24. Esteban O, Birman D, Schaer M, Koyejo OO, Poldrack RA, Gorgolewski KJ. MRIQC: Advancing the automatic prediction of image quality in MRI from unseen sites. PLoS ONE 2017;12:e0184661.

25. Triantafyllou C, Hoge RD, Krueger G, et al. Comparison of physiological noise at 1.5 T, 3 T and 7 T and optimization of fMRI acquisition parameters. NeuroImage 2005;26:243–50.

26. Welvaert M, Rosseel Y. On the Definition of Signal-To-Noise Ratio and Contrast-To-Noise Ratio for fMRI Data. PLoS ONE 2013;8:e77089.

27. Frazier JA, Chiu S, Breeze JL, et al. Structural Brain Magnetic Resonance Imaging of Limbic and Thalamic Volumes in Pediatric Bipolar Disorder. Am J Psychiatry 2005;162:1256–65.

28. Amunts K, Mohlberg H, Bludau S, Zilles K. Julich-Brain: A 3D probabilistic atlas of the human brain’s cytoarchitecture. Science 2020;369:988–92.

29. Esteban O, Ciric R, Finc K, et al. Analysis of task-based functional MRI data preprocessed with fMRIPrep. Nat Protoc 2020;15:2186–202.

30. Gorgolewski K, Burns CD, Madison C, et al. Nipype: A Flexible, Lightweight and Extensible Neuroimaging Data Processing Framework in Python. Front Neuroinform 2011;5:13.

31. Tustison NJ, Avants BB, Cook PA, et al. N4ITK: Improved N3 Bias Correction. IEEE Trans Méd Imaging 2010;29:1310–20.

32. Avants B, Tustison NJ, Song G. Advanced Normalization Tools: V1.0. Insight J 2009.

33. Zhang Y, Brady M, Smith S. Segmentation of brain MR images through a hidden Markov random field model and the expectation-maximization algorithm. IEEE transactions on medical imaging 2001;20:45–57.

34. Smith SM. Fast robust automated brain extraction. Human Brain Mapping 2002;17:143–55.

35. Jenkinson M, Smith S. A global optimisation method for robust affine registration of brain images. Medical Image Analysis 2001;5:143–56.

36. Greve DN, Fischl B. Accurate and robust brain image alignment using boundary-based registration. NeuroImage 2009;48:63–72.

37. Jenkinson M. Improved Optimization for the Robust and Accurate Linear Registration and Motion Correction of Brain Images. NeuroImage 2002;17:825–41.

38. Power JD, Mitra A, Laumann TO, Snyder AZ, Schlaggar BL, Petersen SE. Methods to detect, characterize, and remove motion artifact in resting state fMRI. NeuroImage 2014;84:320–41.

39. Satterthwaite TD, Elliott MA, Gerraty RT, et al. An improved framework for confound regression and filtering for control of motion artifact in the preprocessing of resting-state functional connectivity data. Neuroimage 2013;64:240–56.

40. Lanczos C. A Precision Approximation of the Gamma Function. J Soc Ind Appl Math Ser B Numer Anal 1964;1:86–96.

41. Andersson JLR, Hutton C, Ashburner J, Turner R, Friston K. Modeling Geometric Deformations in EPI Time Series. NeuroImage 2001;13:903–19.

42. Ashburner J, Friston KJ. Unified segmentation. NeuroImage 2005;26:839–51.

43. Andersson J, Jenkinson M, Smith S. Non-linear registration, aka Spatial normalisation. FMRIB technical report TR07JA2 2007.

44. Woolrich MW, Ripley BD, Brady M, Smith SM. Temporal autocorrelation in univariate linear modeling of FMRI data. NeuroImage 2001;14:1370–86.

45. Pruim RHR, Mennes M, Buitelaar JK, Beckmann CF. Evaluation of ICA-AROMA and alternative strategies for motion artifact removal in resting state fMRI. NeuroImage 2015;112:278–87.

46. Beckmann CF, Smith SM. Probabilistic independent component analysis for functional magnetic resonance imaging. Medical Imaging, IEEE Transactions on 2004;23:137–52.

47. Shirer WR, Jiang H, Price CM, Ng B, Greicius MD. Optimization of rs-fMRI Pre-processing for Enhanced Signal-Noise Separation, Test-Retest Reliability, and Group Discrimination. NeuroImage 2015;117:67–79.

48. Fales KR, Zhi X, Song H, Lazar NA. Replicability of Functional Brain Networks: A Study Through the Lens of Seven Resting-State Networks. Hum Brain Mapp 2026;47:e70559.

